# Evaluating new evidence in the early dynamics of the novel coronavirus COVID-19 outbreak in Wuhan, China with real time domestic traffic and potential asymptomatic transmissions

**DOI:** 10.1101/2020.02.15.20023440

**Authors:** Can Zhou

## Abstract

The novel coronavirus (COVID-19), first detected in Wuhan, China in December 2019, has spread to 28 countries/regions with over 43,000 confirmed cases. Much about this outbreak is still unknown. At this early stage of the epidemic, it is important to investigate alternative sources of information to understand its dynamics and spread. With updated real time domestic traffic, this study aims to integrate recent evidence of international evacuees extracted from Wuhan between Jan. 29 and Feb. 2, 2020 to infer the dynamics of the COVD-19 outbreak in Wuhan. In addition, a modified SEIR model was used to evaluate the empirical support for the presence of asymptomatic transmissions. Based on the data examined, this study found little evidence for the presence of asymptomatic transmissions. However, it is still too early to rule out its presence conclusively due to sample size and other limitations. The updated basic reproductive number was found to be 2.12 on average with a 95% credible interval of [2.04, 2.18]. It is smaller than previous estimates probably because the new estimate factors in the social and non-pharmaceutical mitigation implemented in Wuhan through the evacuee dataset. Detailed predictions of infected individuals exported both domestically and internationally were produced. The estimated case confirmation rate has been low but has increased steadily to 23.37% on average. The findings of this study depend on the validity of the underlying assumptions, and continuing work is needed, especially in monitoring the current infection status of Wuhan residents.

## Background

The novel coronavirus (COVID-19) was first detected in Wuhan, China in December 2019, and three months later, 28 countries/regions have reported confirmed cases of COVID-19 infections, with a total of 43,101 confirmed cases globally (22pm CST on Feb. 10, 2020). Illness associated with COVID-19 infection ranges from asymptomatic or mild respiratory illness to severe respiratory symptoms and death (Guan et al., 2020). At the time of this writing, there has been 1,018 confirmed deaths globally from the infection.

Much about this virus is still unknown at this stage (see Figure 1 for a timeline of important events). The initial batch of confirmed cases has links with a live animal market in Wuhan (Huanan seafood market), suggesting a zoonotic origin (Li et al., 2020). It is suspected that the virus originated from bats, but the definite source and its reservoir host(s) are still unknown. Infections among colleagues and family members, who do not have a recent travel history to Wuhan, and also among health care workers confirm the human-to-human transmission (Bastola et al., 2020; Chan et al., 2020; Li et al., 2020; Phan et al., 2020; World Health Organization, 2020). A recent concern is the potential presence of asymptomatic transmission. One confirmed case in Germany suggests that the infection may have been transmitted during the incubation period of the index patient (Rothe et al., 2020).

**Figure 1.**
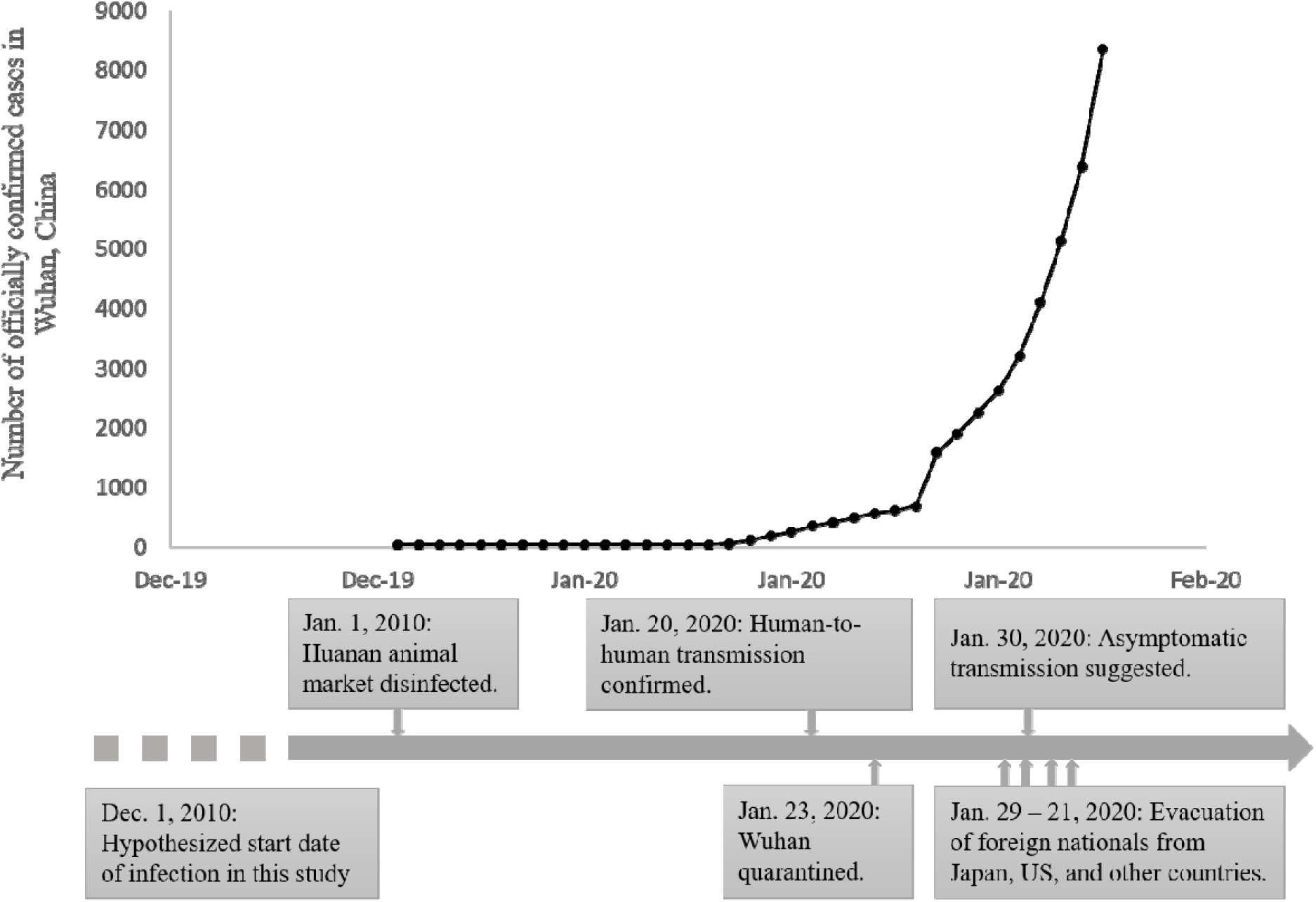
Timeline of important events in the novel coronavirus COVD-19 outbreak and the cumulative number of officially confirmed cases in the city of Wuhan, China.

In the dynamics of an outbreak, the basic reproductive number *R*_*0*_ is an important epidemiological parameter: it measures the number of cases an initial infectious individual can generate in a fully susceptible population. A larger than one *R*_*0*_ indicates the epidemic is growing, while a smaller than one *R*_*0*_ indicates the epidemic is declining. Two separate studies used the Susceptible-Exposed-Infected-Resistant (SEIR) model to study the epidemiological dynamics of the outbreak in Wuhan, and the estimated *R*_*0*_ ranges from 2.68 to 3.8 (Read, Bridgen, Cummings, Ho, & Jewell, 2020; Wu, Leung, & Leung, 2020). The estimate of *R*_*0*_ shows much variation among different studies, and also within each study with different model assumptions. In addition, both studies used different traffic data to model the population flow: historical air travel was used in Read et al. (2020), and historical air, train and road travel was used in Wu et al. (2020). It is assumed that the real time traffic pattern in Wuhan is not affected by the outbreak. However, this assumption needs to be verified with the real time population flow, because Wuhan, as a major air and train traffic hub in the central China and with a 36% floating population, has the capacity to drive substantial day-to-day variation in traffic.

It is important to investigate alternative sources of information to understand the disease dynamics due to the inherent issues associated with the time series of the confirmed cases in China. The daily confirmed cases will likely show an increasing trend purely due to the increase in sample processing throughput. Since Jan. 1, 2020, the Chinese CDC has progressively adopted different versions of the case definition, simplified the procedural burden to confirm a case and subsequently increased the throughput of processing virus samples. One study used a hypothesized temporally increasing confirmation rate to estimate *R*_*0*_ (Zhao et al., 2020). However, the actual confirmation rate is unknown and the estimated *R*_*0*_ is highly variable. Another source of uncertainty is the suspected low confirmation rate of this disease. It has been estimated that only 5.1% of infections in Wuhan are identified (Read et al., 2020). With these concerns, Wu et al. (2020) and Nishiura, Jung, et al. (2020) used an alternative data source, i.e., the number of internationally exported cases between Dec. 31, 2019 and Jan. 28, 2020, to infer the disease dynamics.

Another alternative source of information comes from the evacuated foreign nationals, which provide snapshot views of the status of the epidemic in the city of Wuhan. Between Jan 29 and Feb 2, 2020, there have been 2,666 foreign nationals evacuated out of Wuhan, and their health has been closely monitored after arrival at their destination, 12 of them have been tested positive for COVD-19 infection as of this writing (Feb 11, 2020). There has been at least 9 days between the latest date of evacuation included in this study and the last day of confirmation to avoid any bias in the infection rate. It should be emphasized that these evacuation flights took place after the city of Wuhan was quarantined, and all the successfully evacuated individuals have passed at least two rounds of body temperature screenings at the airport. In this study, it is assumed that these evacuees provide a representative sample of the status of the general population in Wuhan at the time of extraction. A subset of the evacuees data has been analyzed in Nishiura, Kobayashi, et al. (2020)

With updated real time domestic traffic data, this study aims to update the current status of the COVD-19 outbreak in the city of Wuhan with the alternative information from the evacuees between the end of January and the start of February. In addition, the currently used prediction model for this outbreak, i.e., SEIR, does not allow for potential transmission of infection from latent individuals. In this study, the author modifies the SEIR model to evaluate the empirical support for the presence of asymptomatic transmissions based on current evidence.

## Data and methods

### Internationally exported cases

The original internationally exported cases used in Wu et al. (2020) contain 63 cases, of which 4 cases do not have a recent travel history to Wuhan and they were excluded from analysis. Additionally, one confirmed case in Germany infected through contacts with a patient from Wuhan (Rothe et al., 2020), and it is also excluded here. A total of 58 confirmed cases from 14 countries/regions were used in this study. In this study, the author adopts the same case definition as in Wu et al. (2020): a case is an individual that show symptoms, which can be detected by temperature screenings, or severe enough to require hospitalization, plus a recent travel history to Wuhan. In Wu et al. (2020), the symptom onset date was used as the date of exportation. However, the onset date does not always coincide with the date of travel from Wuhan. In this study, the date of exportation is defined as the date the individual exits Wuhan (local time, 49 cases), and when the exit date is not available, the symptom onset date was used (4 cases), and when both the exit date and the onset date were not available, the confirmation date was used instead (4 cases). The export dates fall between Dec. 31, 2019 and Jan. 29, 2019. Of the 40 cases, on which both the date existing Wuhan and the symptom onset date were available, 57.5% (23 cases) were asymptomatic when leaving Wuhan, 25% (10 cases) developed symptoms before leaving Wuhan, and 17.5% (7 cases) developed symptoms on the same date when leaving Wuhan.

### Successfully evacuated foreign nationals

The infection status of a total of 2,666 individuals that successfully evacuated from Wuhan between Jan. 29 and Feb. 2, 2020 were used in this study. These individuals have passed through body temperature screenings at the time exiting Wuhan, and their samples have been analyzed for infection and their health monitored after arrival at their destination. As of Feb. 10, 2020, 12 of these evacuees have been tested positive for COVD-19 infection, and many of those initially tested negative for virus infection are still under quarantine. Based on the case definition adopted in this study, all the infectious individuals are excluded from the evacuees, and it is further assumed that the false positive rate of the body temperature screening is the same across all the individuals. Real time population flow into/out of Wuhan

Every year, Baidu Migration (https://qianxi.baidu.com/) tracks the real time population flow for chunyun, a period with extremely high traffic flow in China around the Lunar New Year, from major Chinese cities based on usage patterns of Baidu services. Baidu Migration offers a daily heat index for the intensity of the population flow both out of and into Wuhan. Here, it is assumed that the heat index is linear with the size of population flow. To estimate the scaling factor *s* between the absolute size of the traffic and the heat index *h*, the historical population flow out of Wuhan *L*_*w,c*_ and into Wuhan *L*_*c,w*_ in 2019 were used,

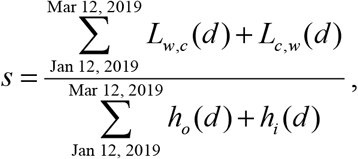

where *h*_*o*_(*d*) and *h*_*i*_(*d*) are respectively the heat index for the traffic moving out of and into Wuhan on day *d*, Jan. 12, 2019 and Mar. 12, 2019 are the end dates when the Baidu Migration data are available in 2019, and they correspond to the period between Jan. 1, 2020 and Feb. 18, 2020 according to Lunar Calendar, *L*_*w,c*_=502,013, *L*_*c,w*_=487,310 before and after 2019 chunyun, and *L*_*w,c*_=717,226, *L*_*c,w*_=810,500 during 2019 chunyun (between Jan. 21 and Mar. 1, 2019) based on Wu et al. (2020). The scaling factor *s* was found to be 138,412.01 individuals per unit of the heat index. It is further assumed that the scaling factor is similar between 2019 and 2020. With the estimated scaling factor *s*, the number of people at time *t* moving out of Wuhan *L*_*w,c*_(*t*) and into Wuhan *L*_*c,w*_(*t*) domestically was calculated based on real time Baidu Migration heat index *h*_*i*_(*t*) at time *t* between Jan. 1, 2020 to Feb 7, 2020

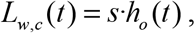

and

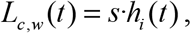

where *h*_*o*_ (*t*) and *h*_*i*_ (*t*) are respectively the heat index for traffic moving out of and into Wuhan at time *t* in 2020.

### Models

An open population SEIR epidemiology model (Figure 2) with a latent period (*D*_*E*_) of 3 days (Guan et al., 2020), infectious period (*D*_*I*_) of 5.4 days and a mean serial interval (*D*_*E*_*+D*_*I*_) of 8.4 days based on SARS (Lipsitch et al., 2003; Wu et al., 2020):

**Figure 2.**
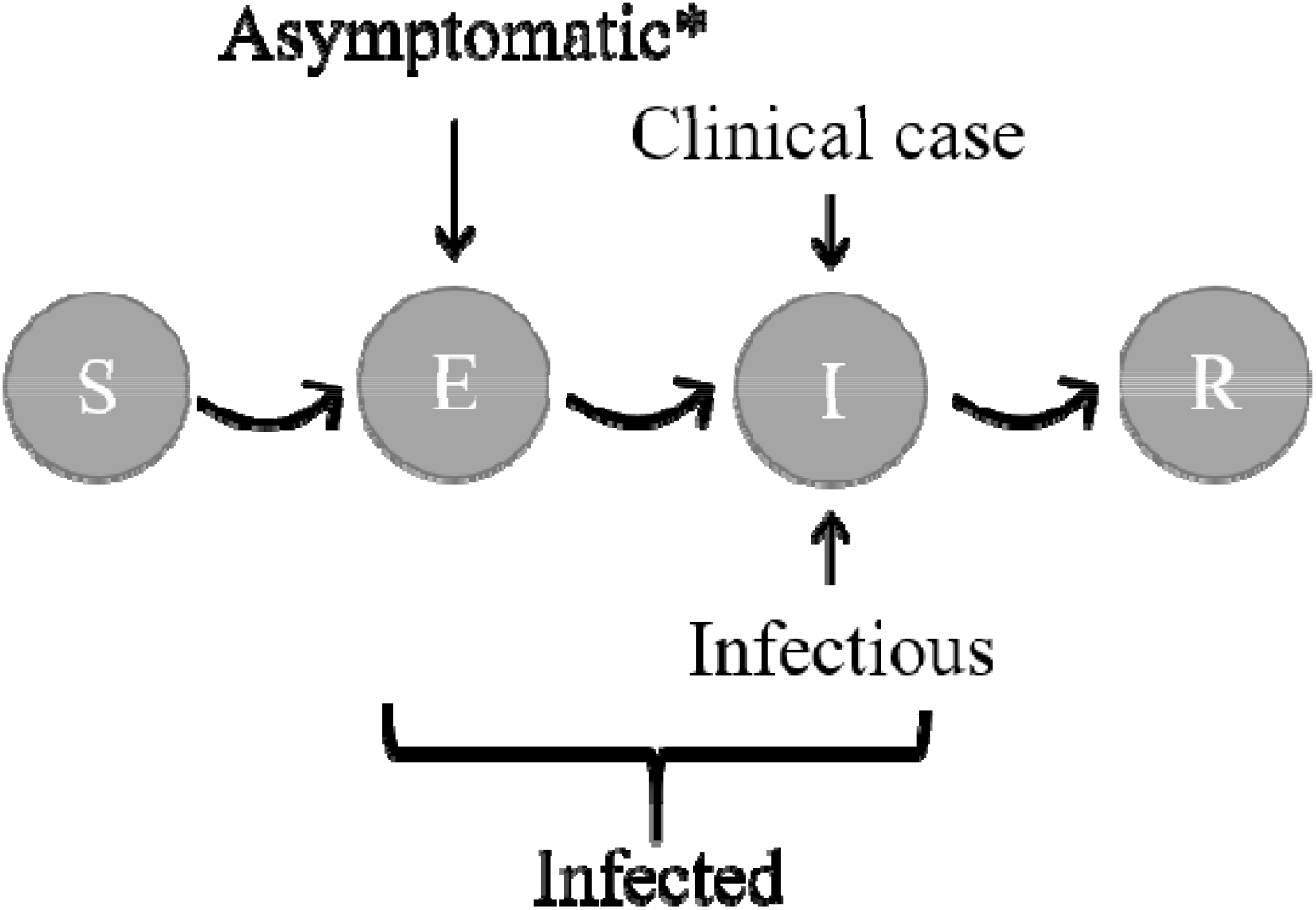
State transition graph of a SEIR model. ^*^Latent individuals do not show any symptoms that can be detected by body temperature screening or are severe enough to require hospitalization, but they are otherwise allowed to exhibit other non-specific minor symptoms.

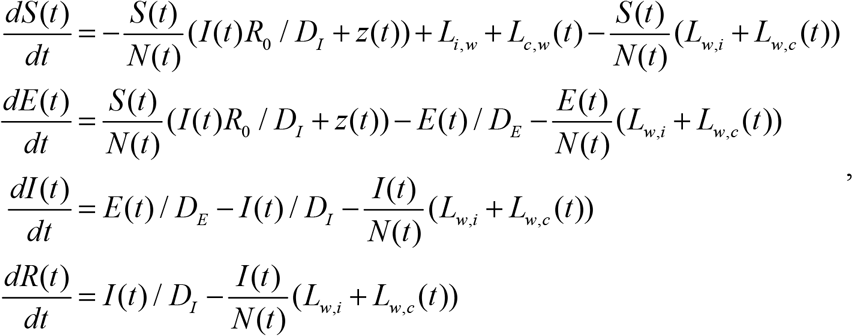

where *S*(*t*), *E*(*t*), *I*(*t*), *R*(*t*) were the number of susceptible, latent, infected and removed individuals in the population at time *t*, respectively, *N*(*t*) is the total number of individuals in the population, *R*_*0*_ is the basic reproductive number, *L*_*ij*_(t) is the daily number of individuals migrating from *i* to *j*, for *i, j* ∈{*w*: Wuhan, *i*: international, *c*: other cities of China}, and *z*(*t*) is the daily force of zoonotic infection. It is assumed that the dynamics start on Dec. 1, 2019 (day 0) with a population of *c*. 14 million susceptible individuals, initiated with a constant zoonotic infection equivalent to 43×2 infectious individuals until Jan. 1, 2020 (day 31) when the Huanan seafood market was disinfected. In this study, we only model the disease dynamics in the city of Wuhan, which has 9.08 million permanent residents and 5.10 million floating population according to the most recent census data at the end of 2019. The treatment of the strength and duration of the zoonotic infection is the same as the prediction model in Wu et al. (2020). Real time domestic traffic (*L*_*w,c*_ and *L*_*c,w*_) are estimated based on the Baidu Migration heat index and scaling factor *s*; international traffic (*L*_*w,i*_ and *L*_*i,w*_) is based on averaged historical patterns as in Wu et al. (2020). In the baseline model, only one parameter was estimated, i.e., *R*_*0*_, and it was given a non-informative prior *R*_*0*_ ∼ Uniform(0, 10).

To investigate whether the observed infection data support the presence of asymptomatic transmission, an open population SEAIR model (Figure 3) was formulated, which has one additional stage *A*, the asymptomatic infectious stage, between the latent (*E*) and the infectious (*I*) stages of a SEIR model:

**Figure 3.**
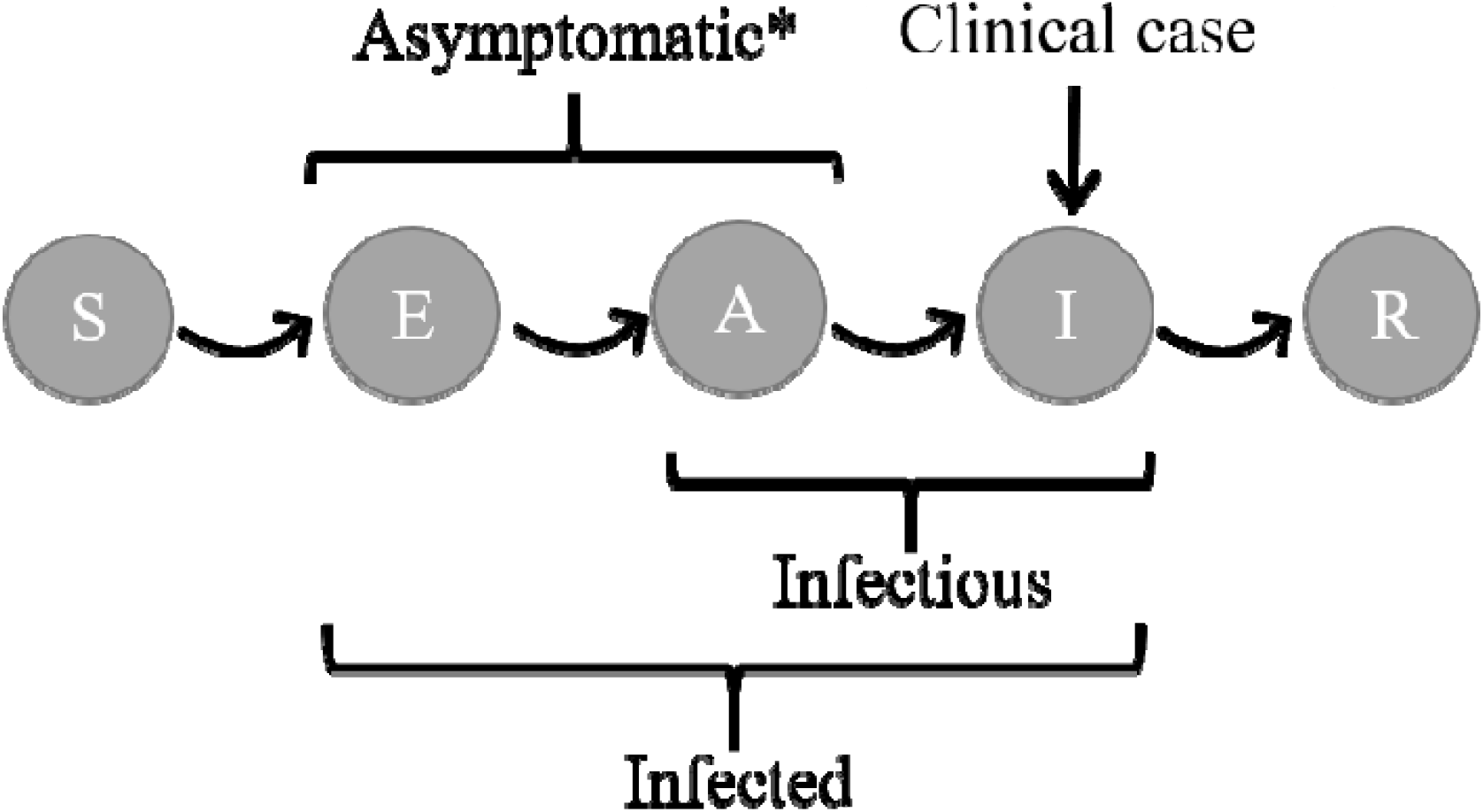
State transition graph of a SEAIR model. Compared to a SEIR model, a SEAIR model has an additional state A that is both asymptomatic and infectious. ^*^An asymptomatic individual does not show any symptoms that can be detected by body temperature screening or are severe enough to require hospitalization, but they are otherwise allowed to exhibit other non-specific minor symptoms.

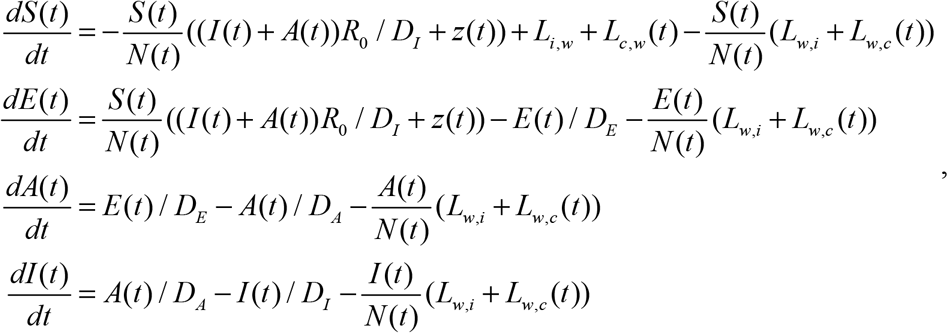

where *R*_*0*_ is the basic reproductive number of this model, *γ* has a non-informative prior *γ* ∼ Uniform(0,1), and the equation for the removed stage stay unchanged. In this model, the latent period (*D*_*E*_) can be shorter than the mean latent period (3 days) between 0 and 3 days, the asymptomatic infectious period (*D*_*A*_) is (3-*D*_*E*_) days, and the symptomatic infectious period (*D*_*I*_) stays unchanged.

Additional equations were solved alongside the dynamical equations above. These additional equations do not influence the dynamics of the system; they only keep track of variables of interest, such as the number of exported cases internally and domestically, and the number of cases and infected individuals in Wuhan. The cumulative number of exported infected individuals internationally *C*_*i*_(*t*) at time *t* since Dec. 1, 2019 is

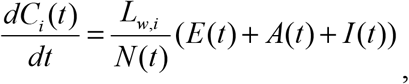

where *A*(*t*) is the number of asymptomatic infectious individuals at time *t* in the SEAIR model and set to 0 for all *t* in the SEIR model, and the number of infected individuals exported internationally in day *d* is thus *C*_*i*_(*d*)-*C*_*i*_(*d*-1). Similarly, the cumulative number of infected individuals exported domestically *C*_*c*_(*t*) at time *t* since Dec. 1, 2019 follows

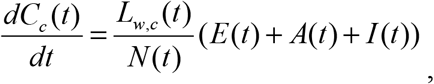

and the number of infected individuals exported domestically in day *d* is thus *C*_*c*_(*d*)-*C*_*c*_(*d*-1). The cumulative number of infected individuals in Wuhan *C*_*w*_(*t*) at time *t* follows

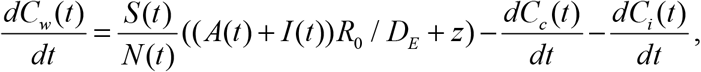

and the number of infected individuals in Wuhan in day *d* is thus *C*_*w*_(*d*)-*C*_*w*_(*d*-1). The cumulative number of clinical cases in Wuhan *C*_*I*_(*t*) at time *t* follows

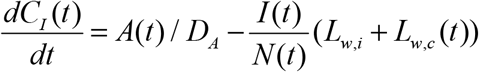

for the SEAIR model, and

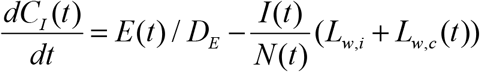

for the SEIR model.

It is assumed that the number of observed cases exported internationally *y*(*d*) on date *d* follows a Poisson distribution

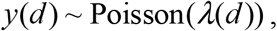

where *λ*_*d*_ = *C*_*i*_ (*d*) − *C*_*i*_ (*d* −1). The number of infected evacuees without symptoms *z*(*d*) (latent and asymptomatic infectious individuals) within a sample of *n*(*d*) evacuees without symptoms (susceptible, latent, asymptomatic infectious and removed individuals) extracted from Wuhan on day *d* is binomial distributed

*z*(*d*) ∼ Binomial(*n*(*d*), *p*(*d*)),

where *p*(*d*) = (*E*(*d*) + *A*(*d*)) / (*S* (*d*) + *E*(*d*) + *A*(*d*) + *R*(*d*)), and in the SEAR model, *A*(*d*) is universally 0.

Internationally exported cases alone does not offer inference into parameter *γ* because the number of individuals from all the infected stages (stages *E, A* and *I* in the SEAIR model) are mixed together in the likelihood function and thus not separable. On the other hand, the evacuee dataset allows us to separate asymptomatic individuals (*A* and *I*) from the rest because of the body temperature screenings at the airport. With both datasets, it is thus possible to check if the observed infection data support a shortened non-infectious latent period, i.e., evidence for the existence of an asymptomatic infectious period (*A*).

### Model estimation and evaluation

The ordinary differential equations were solved using the LSODA algorithm (Petzold & Hindmarsh, 1997) with both the absolute and relative tolerance set to 10^−3^. Multiple chains with a Metropolis-Hastings sequential sampler were used to draw samples from the posterior of the model parameters. Python packages numpy and pymc3 were used to estimate model parameters, and all the data processing was conducted in Python 3.7 (Rossum, 1995) and statistical program R 3.6 (R Development Core Team, 2016). All the datasets used in this study are hosted on a public repository and made available at https://github.com/HVoltBb/2019nCov.

Model performance was evaluated using the WAIC (widely applicable information criterion) (Gelman, Hwang, & Vehtari, 2014), calculated as

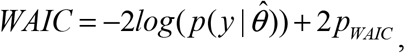

where 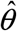 is the posterior mean estimate, and *p*_*WAIC*_ is the data based bias correction term. The model with a smaller WAIC has better predictive performance than a model with a larger WAIC. The ΔWAIC_*i*_ for model *i* was calculated as WAIC_*i*_ − *min*(WAIC), where the minimum is taken across all the candidate models.

The empirical support of each candidate model can be evaluated using WAIC weights *w*_*i*_, which takes values between 0 and 1, for *i*=1, …, *M*,

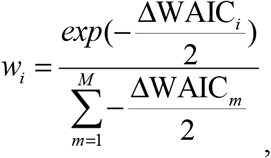

where *M* is the number of candidate models. Models with larger weights have more support from the empirical data than models with smaller weights. These weights can be used to construct an ensemble estimator *ĥ*,

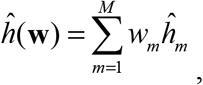

where *ĥ*_*m*_ is the estimator from candidate model *m*. The major advantage of using the ensemble estimator is its ability to account for model uncertainty and obtain better predictive performance than any single constituent candidate model.

To assess the significance of the SEAIR model with respect to the SEIR model, parametric bootstrap samples were generated to estimate the small sample distribution of the model weights when the data generating model is SEIR. The simulation procedure is as follows. First, a random sample was drawn from the posterior of the *R*_*0*_ in model SEIR, the SEIR model was solved for each of these *R*_*0*_s, then the number of internationally exported cases simulated from the Poisson distribution for all the dates when such data are available, and the number of latent individuals simulated from the Binomial distribution for each sample of evacuated foreign nationals. Next, for each simulated data set, posterior estimates were drawn from both the SEIR and SEAIR models, and WAIC and weight *w* calculated for both models. The significance of the weight of the SEAIR model is evaluated against the simulated empirical null distribution of the weight when the true model is SEIR.

## Results

Based on rescaled real time domestic migration data, the population outflow from the city of Wuhan differed significantly from its historical pattern in 2019. The net outflow between the day the first COVD-19 case was confirmed and when Wuhan was quarantined shows an exponential increase pattern (Figure 4). During this period, the overall outflow was 15% more than the same period (according to Lunar Calendar) in 2019. At its peak (days before Wuhan was quarantined), over millions of individuals were leaving Wuhan on a daily basis. Two days after Wuhan was quarantined, the net outflow reserved signs and stays at a relatively low level.

**Figure 4.**
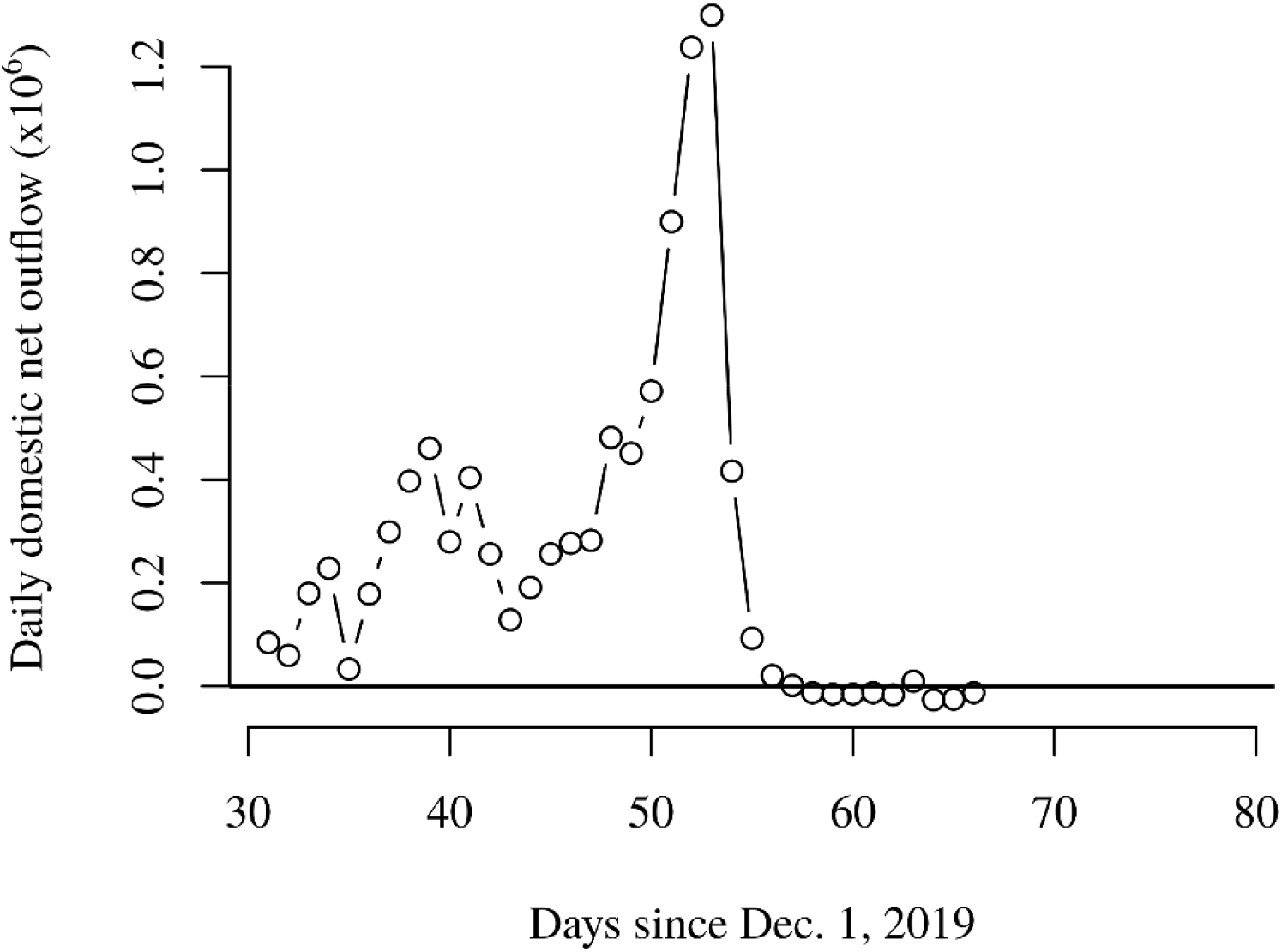
Estimated real time domestic population net outflow from the city of Wuhan, China based on Baidu Migration calibrated by historical Tencent mobility and OAG data. The sharp decline in traffic on day 54 since Dec. 1, 2019 (Feb. 23, 2020) corresponds to the day when the city of Wuhan was quarantined.

Both the SEIR and SEAIR models were fitted to the infection data, and SEIR model has better performance than the SEAIR model based on WAIC (Table 1). However, the difference in WAIC between both candidate models is small, i.e., 3.12. Based on the estimated null distribution of the Δ WAIC from parametric bootstrap samples, the *p-value* for ΔWAIC = 3.12 is *c*. 0.06, slightly larger than the 5% significance level. Technically, the SEAIR model is not statistically significant at the 5% level; however, due to the relatively small sample size of the infection data with respect to the number of residents in Wuhan, it is inconclusive whether the disease dynamics show evidence for the SEAIR model. In the following, parameter estimates from both the SEIR and SEAIR models are presented.

**Table 1.** Candidate models and model selection results.

| Model | WAIC | ΔWAIC | $w_{WAIC}$ |
| --- | --- | --- | --- |
| SEIR | 182.24 | 0 | 82.64% |
| SEAIR | 185.36 | 3.12 | 17.36% |

The SEIR model produced an *R*_*0*_ with a mean of 2.12 and a 95% credible interval of [2.04, 2.18] (Figure 5). In the SEAIR model, the estimated *γ* and *R*_*0*_ are highly correlated (Figure 6). The joint posterior distribution obtained the highest density in the upper right corner of their range where *γ* is close to 1 and *R*_*0*_ is around 2.05. When *γ* = 1, the SEAIR model is equivalent to the SEIR model. This result further supports the better performance of the SEIR model for the current infection data, and the lack of evidence for the presence of an asymptomatic infectious stage.

**Figure 5.**
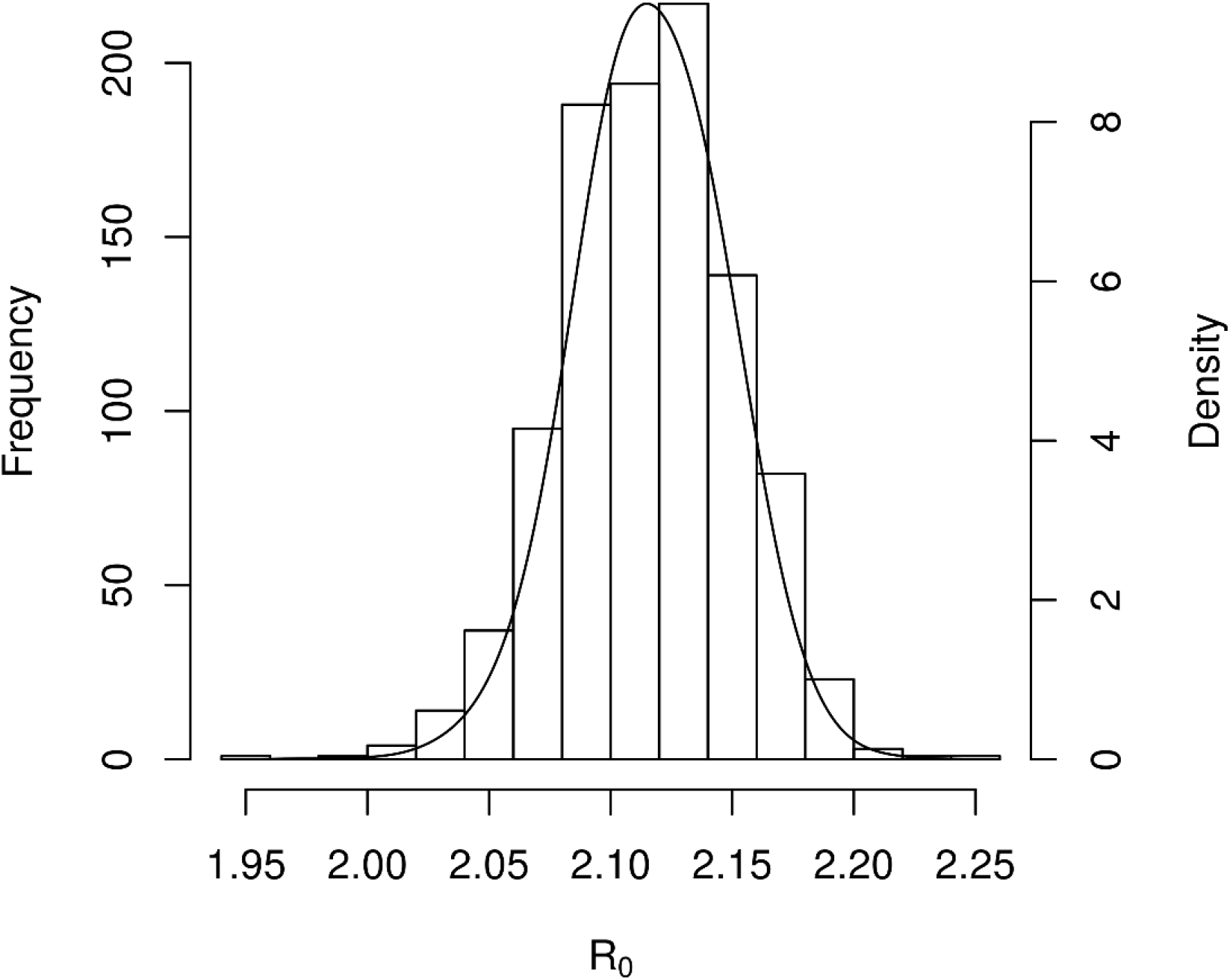
Posterior distribution of the basic reproductive number based on the SEIR model.

**Figure 6.**
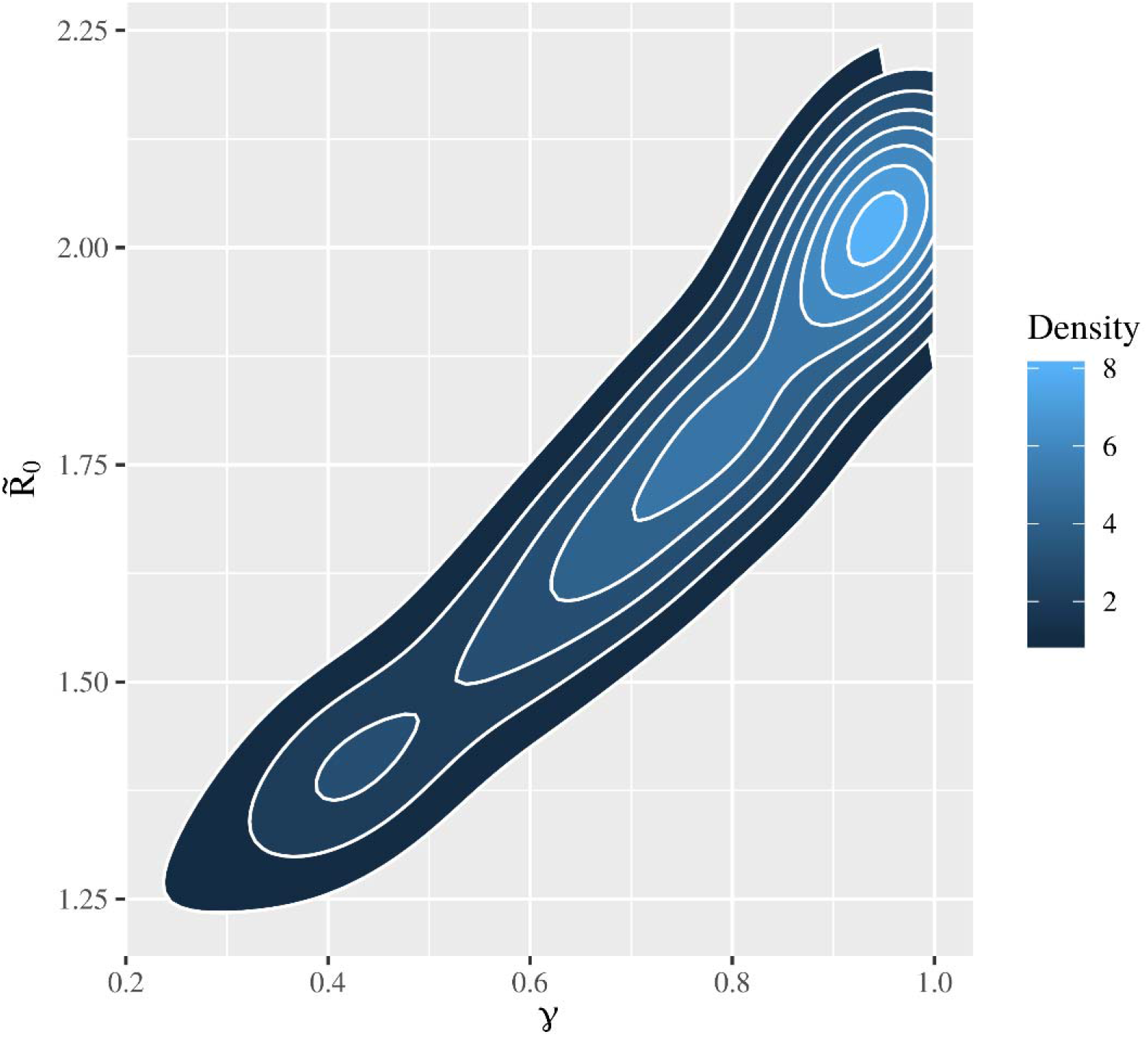
Density plot of the posterior distribution of γ and 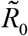 in the SEAIR model.

To avoid the potential danger of omitting asymptomatic transmissions, ensemble model predictions were calculated based on the weighted predictions from both SEIR and SEAIR models. The number of exported infected individuals internationally is approximately linear since day 31 (Jan. 1, 2020) until Wuhan was quarantined (Figure 7). The total number of cases exported internationally is estimated at 146 individuals on average with a 95% credible interval of [121, 173] individuals. The number of infected individuals exported domestically shows an exponentially increasing pattern until Wuhan was quarantined (Figure 8). At its peak, an average of 3,354 infected individuals with a 95% credible interval of [2696, 3996] individuals were exported daily into other cities of China. From Dec. 1, 2019 to Feb. 7, 2020 (the most recent date of Baidu migration data used in this study), the total number of infected individuals exported domestically is estimated to be on average 37,115 individuals with a 95% credible interval of [31354, 43099] individuals.

**Figure 7.**
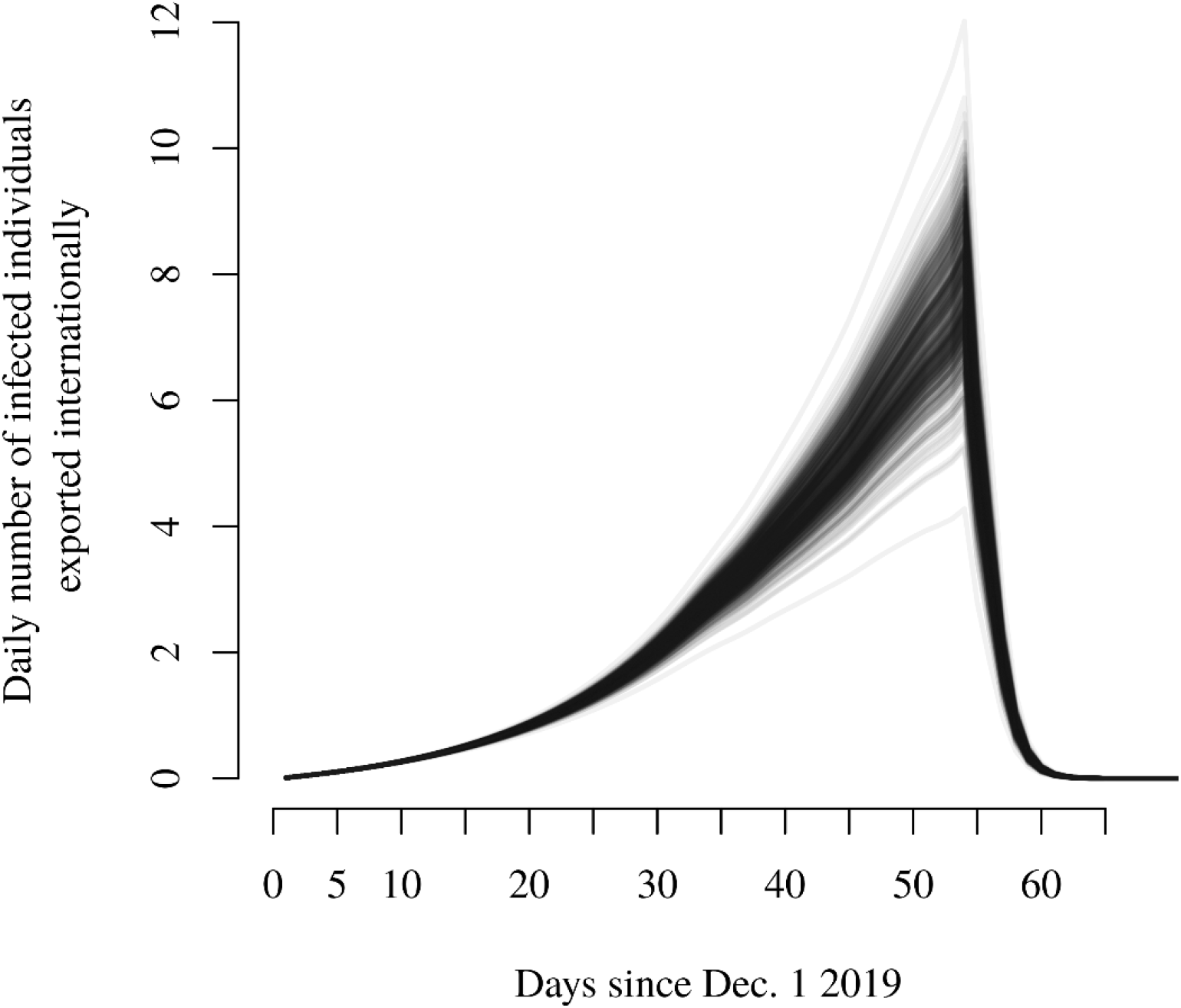
Estimated daily number of infected individuals exported internationally from Wuhan, China. Each curve represents one solution of the posterior estimate.

**Figure 8.**
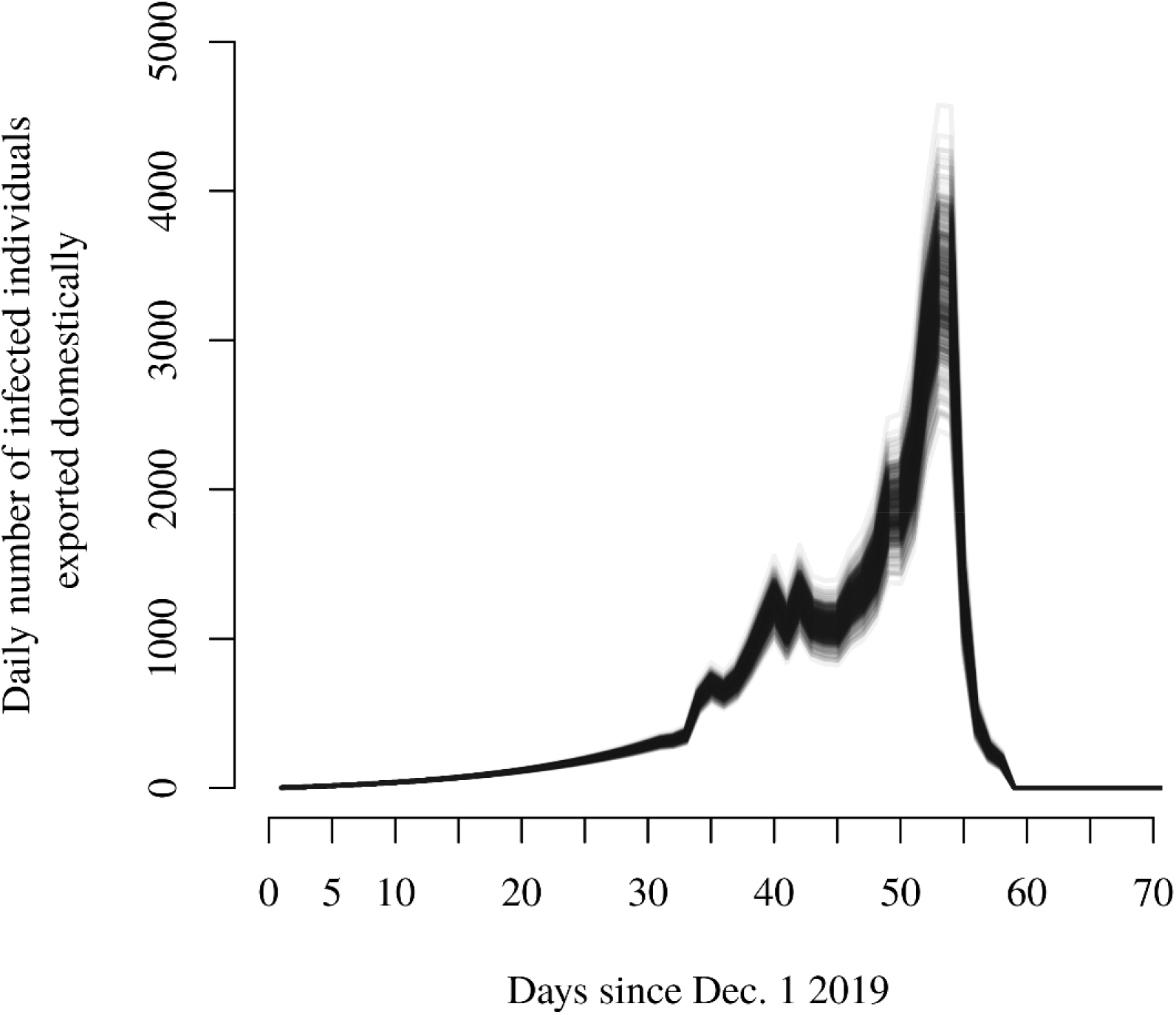
Estimated daily number of infected individuals exported domestically from Wuhan, China. Each curve represents one solution of the posterior estimate.

The cumulative number of infected individuals in Wuhan on Feb. 7, 2020 is estimated at 80,084 on average with a 95% credible interval of [63060, 98567] individuals. Due to the elevated population outflow since Jan. 1, 2020, the number of cumulative infected individuals in Wuhan is approximately linear between day 31 and day 51, then appears to stabilize afterwards, and then increases exponentially after the date Wuhan was quarantined (Figure 9). The predicted pattern closely resembles the trend of the cumulative number of officially confirmed cases in Wuhan with a time lag of approximately 3 days (Figure 1).

**Figure 9.**
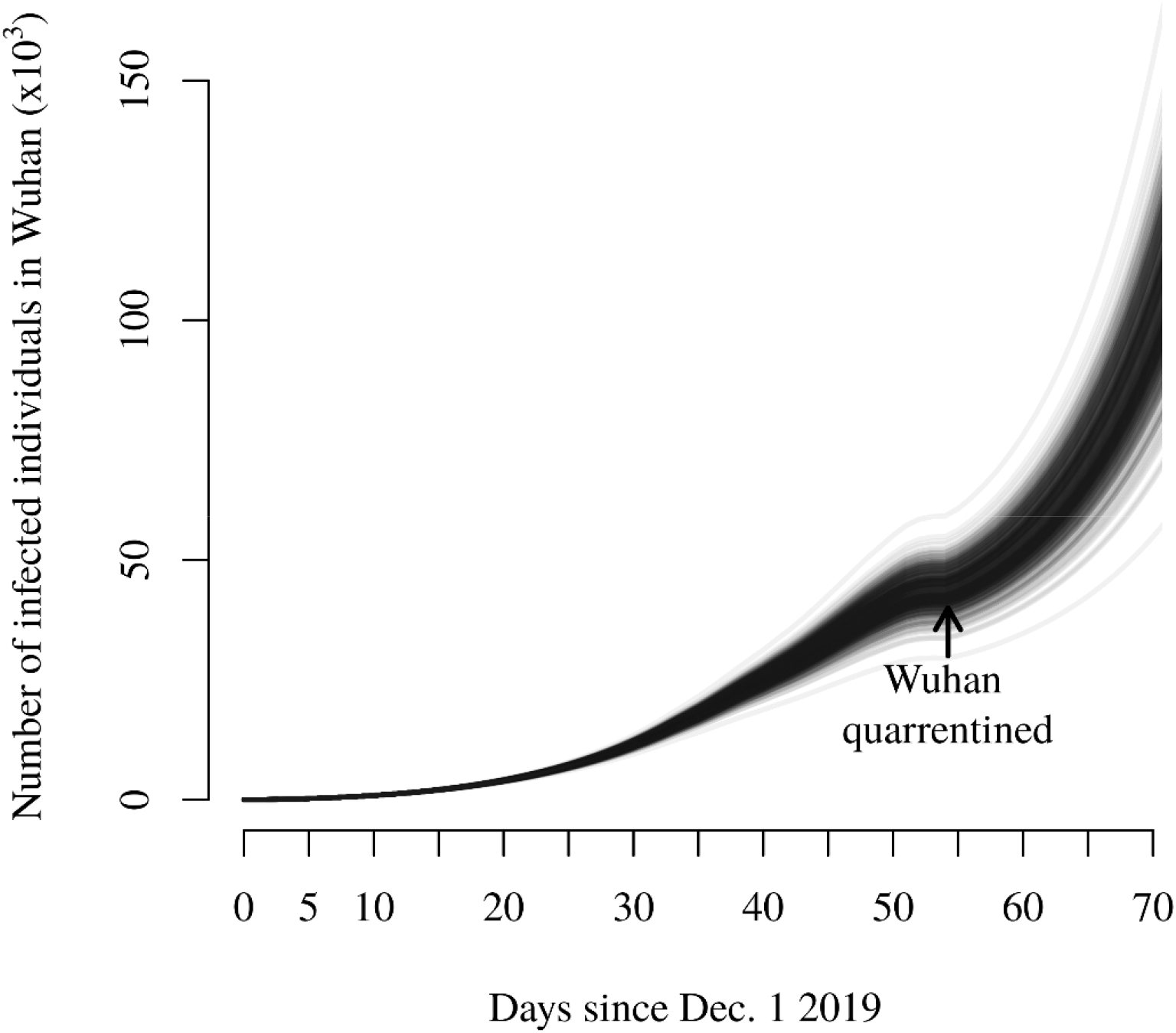
Estimated daily number of infected individuals in Wuhan until Feb. 7, 2019 since Dec. 1, 2019. Each curve represents one solution of the posterior estimate.

Assuming a constant time lag of 3 days, the confirmation rate of this disease has been low, i.e., less than 10%, in January 2019 but has steadily increased ever since (Figure 10). On Feb. 10, 2020, the overall case confirmation rate is estimated at 23.37% on average with a 95% credible interval of [18.72%, 29.26%]. The time lag between the onset of symptoms and official confirmation of the case probably have decreased with time due to people’s increased awareness of this disease and also the reduced procedural burden to confirm a case with newer versions of the case definition from the Chinese CDC. To evaluate other time lags, the estimated case confirmation rate was calculated with lags between 1 and 5 days. These time lags produced similar results on the case confirmation rate on Feb. 10, 2020 (Table 2).

**Table 2.** Mean and lower and upper bounds of the 95% credible interval of the case confirmation rate (%) on Feb. 10, 2010 in Wuhan, China with different time lags between the onset of symptoms and case confirmation.

| Time lag | Mean | Lower bound | Upper bound |
| --- | --- | --- | --- |
| 1 | 20.42 | 16.14 | 25.89 |
| 2 | 21.87 | 17.41 | 27.56 |
| 3 | 23.37 | 18.72 | 29.26 |
| 4 | 24.9 | 20.10 | 31.00 |
| 5 | 26.47 | 21.47 | 32.75 |

**Figure 10.**
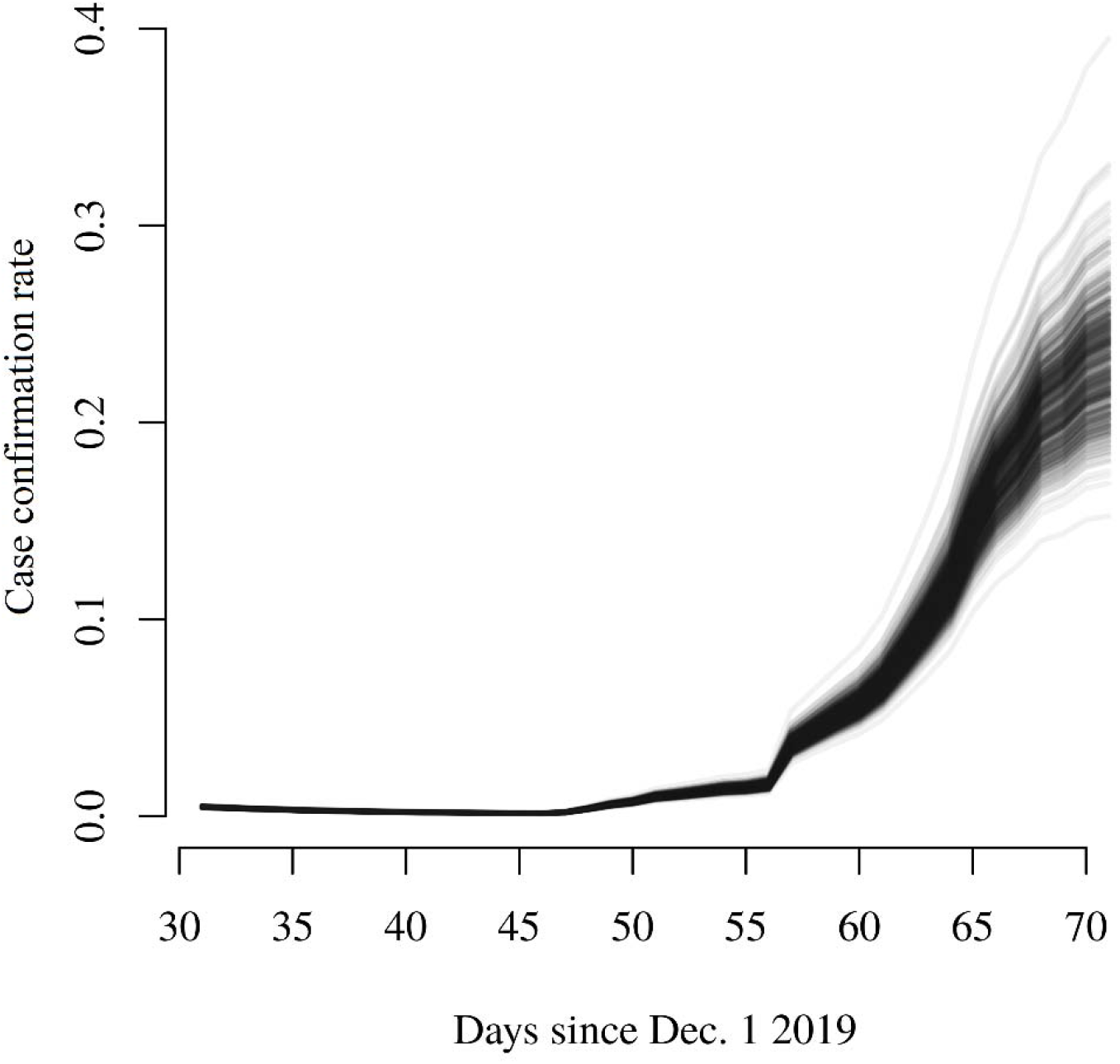
Estimated case confirmation rate. Each curve represents one solution from the posterior estimate.

## Discussions and conclusion

At this stage, our understanding of the dynamics of COVD-19 is still limited due to the relatively low case confirmation, a lack of comprehensive monitoring programs and other limitations. It is thus important to consider alternative information to update our understanding about the outbreak. This study integrates the recent infection data on the early dynamics of the COVD-19 outbreak in Wuhan, China with updated real time domestic traffic data. Little empirical evidence was found supporting the presence of asymptomatic transmission. However, it is still too early to rule out its presence conclusively due to the relatively low sample size and the validity of the assumptions used in this study. The updated basic reproductive number was found to be 2.12 on average, which is significantly smaller than previous estimates (Read et al., 2020; Wu et al., 2020; Zhao et al., 2020), probably because the new estimate factors in the social and non-pharmaceutical mitigation implemented in Wuhan through the evacuee dataset.

The real time traffic is a crucial piece of information to understand the dynamics and the spread of the COVD-19 outbreak in Wuhan. As a major traffic hub in central China, Wuhan’s capacity to drive significant population flows is driving the accelerated spread of the disease domestically. Real time traffic flow out of Wuhan was found to differ significantly from its historical patterns. From Jan. 1, 2020, when the Huanan seafood market was disinfected, to Jan. 23, 2020, when the city of Wuhan was quarantined, the net traffic flow has always been outwards. During this period, the overall net outflow is estimated to be over 9 million individuals. On Feb. 11, 2020, the disease has already spread to all provinces of China with 44,653 confirmed cases in China. In early January, the number of cases in Wuhan is only growing linearly (Figure 9) and even appears to have stabilized days before Jan. 23, due to the accelerated outflow. It is thus very dangerous to overinterpret the temporal pattern of the number of cases without taking into account real time traffic.

A limitation of this study is that the international traffic still relies on its historical patterns in 2019. Due to the elevated domestic outflow, it is reasonable to expect a similar elevated outflow in the international traffic. The internationally exported cases inform the disease dynamics through the proportion of infected outbound international travelers from Wuhan. Increasing the outward flow effectively reduces the proportion infected and subsequently the basic reproductive number. Generally, international traffic by air is less variable than domestic traffic by road, and the potential surge in outbound international traffic would be less than 15% (the surge in outbound domestic traffic). With a 15% increase in the international outbound flow, sensitivity analysis shows a *c*. 2% decrease in the estimated basic reproductive number. Thus, the major conclusions of this study would not be affected by a potential surge. On the other hand, it is still important to update the model and verify the underlying assumptions when the real time international air traffic data become available.

Because of the inherent bias in the daily traffic data, this study refrains from predicting the disease dynamics in other cities of China using meta-population modeling, even though daily real time traffic data among cities are available. The transportation pattern in China is unique in that primary modes of travel is by road and train, which were expected to account for 96% of the holiday traffic in 2020 (Xinhua News Agency, 2020). Many of those trips would take multiple days. For example, a trip from Wuhan to Beijing would span days if the traveler chooses to travel by road. But the real time traffic data would erroneously indicate flow from Wuhan into some traffic hubs in between Wuhan and the final destination, because of the daily accounting unit used in the calculation of the flow pattern. Thus, a meta-population model with such traffic data would substantially over-estimate the imported cases into major traffic hubs around Wuhan, and subsequently under-estimate the imported cases to more distant, but popular destination cities, such as Wenzhou. Air travel on the other hand is less affected by this issue. To solve this problem, the original user usage data collected by Tencent or Baidu need to be reprocessed to reveal those direct long-distance trips.

It is suggested that the first confirmed case in Germany is the result of an asymptomatic transmission, although the index patient at that time did experience some minor non-specific symptoms (Rothe et al., 2020). From December, 2019 to early January, 2020, the estimated mean incubation period is 6.1 days (Li et al., 2020); however, a more recent study put the median incubation period at 3 days (Guan et al., 2020). The heightened awareness of the disease from both the general public and healthcare workers may play a role in discovering early symptoms of this disease. In this study, the estimated most probable asymptomatic infectious period is close to zero and offers little evidence for the presence of asymptomatic transmission in the disease dynamics.

The findings of this study depend on the validity of the underlying assumptions, all of which may render the model inference invalid, and continuing work is needed, especially in monitoring the current status of residents in Wuhan. As shown previously, the assumptions on the start date and the infectious force of the zoonotic infection influences the inference on the basic reproductive number (Read et al., 2020; Wu et al., 2020). Reducing the zoonotic infection force or shortening its duration has the effect of increasing the estimate of the basic reproductive number, and vice versa. In addition, all the infection data used in this study are with foreign nationals, with the assumption that they have a similar virus transmissibility with the Chinese residents. However, it is known that the basic reproductive number vary with human social behavior and organization (Delamater, Street, Leslie, Yang, & Jacobsen, 2019). Therefore, it is important to verify this assumption with local monitoring data.

## Data Availability

Data used in this study are publicly available at the author's GitHub repository.

https://github.com/HVoltBb/2019nCov

## Acknowledgments

The author is thankful to Dr. X.G. for providing insights into and firsthand experience of the COVD-19 outbreak in Wuhan, and Dr. M.F. for reviewing an earlier version of this manuscript. The author also thanks Baidu Migration for providing open access to the real time domestic traffic data. The scientific results and conclusions, as well as any views or opinions expressed herein, are those of the author’s and do not necessarily reflect those of the hosting institution(s). The author declares no conflict of interest for this study.

